# Do Face Masks Create a False Sense of Security? A COVID-19 Dilemma

**DOI:** 10.1101/2020.05.23.20111302

**Authors:** Youpei Yan, Jude Bayham, Eli P. Fenichel, Aaron Richter

## Abstract

Face masks have become an emblem of the public response to COVID-19, with many governments mandating their use in public spaces. The logic is that face masks are low cost and might help prevent some transmission. However, from the start, the assumption that face masks are “low cost” was questioned. Early on, there were warnings of the opportunity cost of public use of medical masks given shortages of personal protective equipment for healthcare providers. This led to recommendations for cloth masks and other face coverings, with little evidence of their ability to prevent transmission. However, there may also be a high cost to these recommendations if people rely on face masks in place of other more effective ways to break transmission, such as staying home. We use SafeGraph smart device location data to show that the representative American in states that have face mask mandates spent 20-30 minutes less time at home, and increase visits to a number of commercial locations, following the mandate. Since the reproductive rate of SAR-COV2, the pathogen that causes COVID-19 is hovering right around one, such substitution behavior could be the difference between controlling the epidemic and a resurgence of cases.

**Highlights:**

- We use smart device location data to show the behavioral response to face mask mandates during the 2020 COVID-19 epidemic.
- We find face mask mandates lead people to spend 20-30 minutes less time at home per day.
- We find face mask mandates increase trip taking to a variety of locations, chief among them are restaurants.
- This substitution behavior is concerning given the limited information on the protective value of casual face coverings.

## Introduction

At the time of writing, 36 States in the United States are requiring some use of face masks in public to combat the COVID-19 epidemic. Similar regulations exist around the world. The evidence that mask-wearing by the general public reduces COVID-19 cases is scant, and the evidence that does exist suggests that the benefits depend on correct usage (Bae et al. 2020; Leung et al. 2020; Mahase 2020; Howard et al. 2020; Mueller and Fernandez 2020). The arguments in favor of masks are that they do no harm, are low cost, and they could provide benefits (Brosseau and Sietsema 2020). However, whether encouraging people to wear masks does no harm and whether they are low cost are important questions. Most recommendations are for cloth masks and non-medical masks because of concerns that the opportunity cost of masks might be high, given the scarcity of masks for medical personnel (Feng et al. 2020). However, there is the potential for even more insidious harm. If individuals substitute mask wearing for other protective behaviors such as allocating more time to staying home, then this false confidence could lead to disinhibition behavior and an increase in COVID-19 cases, a very serious cost. If people who would otherwise stay home and engage in social distancing cease that behavior because they wear masks, then recommendations to wear masks in public may be creating a perverse incentive. Others have pointed out that partially effective vaccines can lead to increased infection, especially given assortative mixing patterns (Talamàs and Vohra 2020). We should expect a similar phenomenon for face masks. Furthermore, messaging around wearing masks could create a false impression of safety.

Substituting the wearing of a face mask for staying home may increase system-wide risk. If the purpose of wearing face masks were only to protect the wearer from COVID-19, then going out with a mask could still be privately optimal so long as masks provide protection equivalent to additional minutes at home. Of course, if people overestimate the private protection conveyed by face masks, then this could be a privately suboptimal decision. It is unlikely, however, that individuals fully account for the costs of their own illness in terms of hospital congestion and other externalized costs. More importantly, the real benefit of mask-wearing or staying home is preventing the spread of COVID-19 by infectious and potentially asymptomatic individuals. Therefore, wearing a mask could create an un-internalized cost if masks do not prevent asymptomatic individuals from spreading the pathogen. Just as wearing a seat belt can encourage less safe driving while conferring no protection to others from such actions, if masks do not prevent infectious individuals from spreading the pathogen, then they induce an externality (Richens, Imrie, and Copas 2000). This is a particular concern, because of the oft cited externality associated with pathogen spread and protective behavior – people typically fail to account for how their behavior prevents spreading the pathogen to others (Gersovitz 2011). The economic theory of social distancing suggests that if people can mitigate disease risks by lower private cost means, then they will distance less (Fenichel 2013).

The concern of inducing risky behaviors by providing some protection is well established in the broad literature on behavior, risk, and externalities. The medical literature uses the term “risk compensation” to describe the case when someone increases certain risky behavior when using protective equipment (Cassell et al. 2006). A close analog is condom use and HIV transmission. In the case of risk compensation, people using condoms engage in more sexual activity and increase the risk to susceptible individuals in the population (Richens, Imrie, and Copas 2000; Shelton 2007). Public health researchers and economists have long been concerned about the behavioral impacts of introducing partially effective prophylaxis or vaccine for viruses such as HIV (Auld 2003; Chen 2006). There exist several examples from economics as well. The potential of endangered species to arrive on one’s land may encourage a landowner to eliminate that species habitat to avoid future land use restrictions (Langpap and Wu 2017). Concerns about energy efficiency and creating a rebound and backfire that creates greater net energy use are also common (Gillingham, Rapson, and Wagner 2016). Pfeiffer and Lin (2014) show how improved irrigation can lead to greater water use.

We contribute to the economics and public health literatures addressing COVID-19 by using the variation in face mask mandates along with mobile device data to measure the change in the amount of time Americans stayed at home, and the number of visits Americans made to public places following face mask mandates. Time at home fell, and visits to public places rose. This is likely risk compensating behavior because face masks are of questionable effectiveness when used in the general population. Though the ultimate impact for face mask orders on transmission also depends on the unknown relative effectiveness in breaking transmission of face masks and staying home.

Well-intentioned public policies may have inherently sent the message that it is safe to go into public so long as one has a mask rather than if one must go out in public a mask might help a bit. Our findings do not suggest policymakers should tell people to stop wearing masks. Rather, policymakers need to be concerned with behavioral feedbacks and need to take time to craft clear messages.

## Materials and Methods

As of writing, there were 36 states mandating face mask use by employees in public-facing businesses, and 16 states ordering all individuals in public spaces to wear face masks (Raifman 2020) (see Figure 1). These orders took effect on the same date for all states except Connecticut (business use on April 3, and all individuals in public on April 20) and Delaware (business use on May 1, and all individuals in public on April 28). For Connecticut and Delaware, we use the first order as the date a face mask order went into effect.

**Figure 1.**
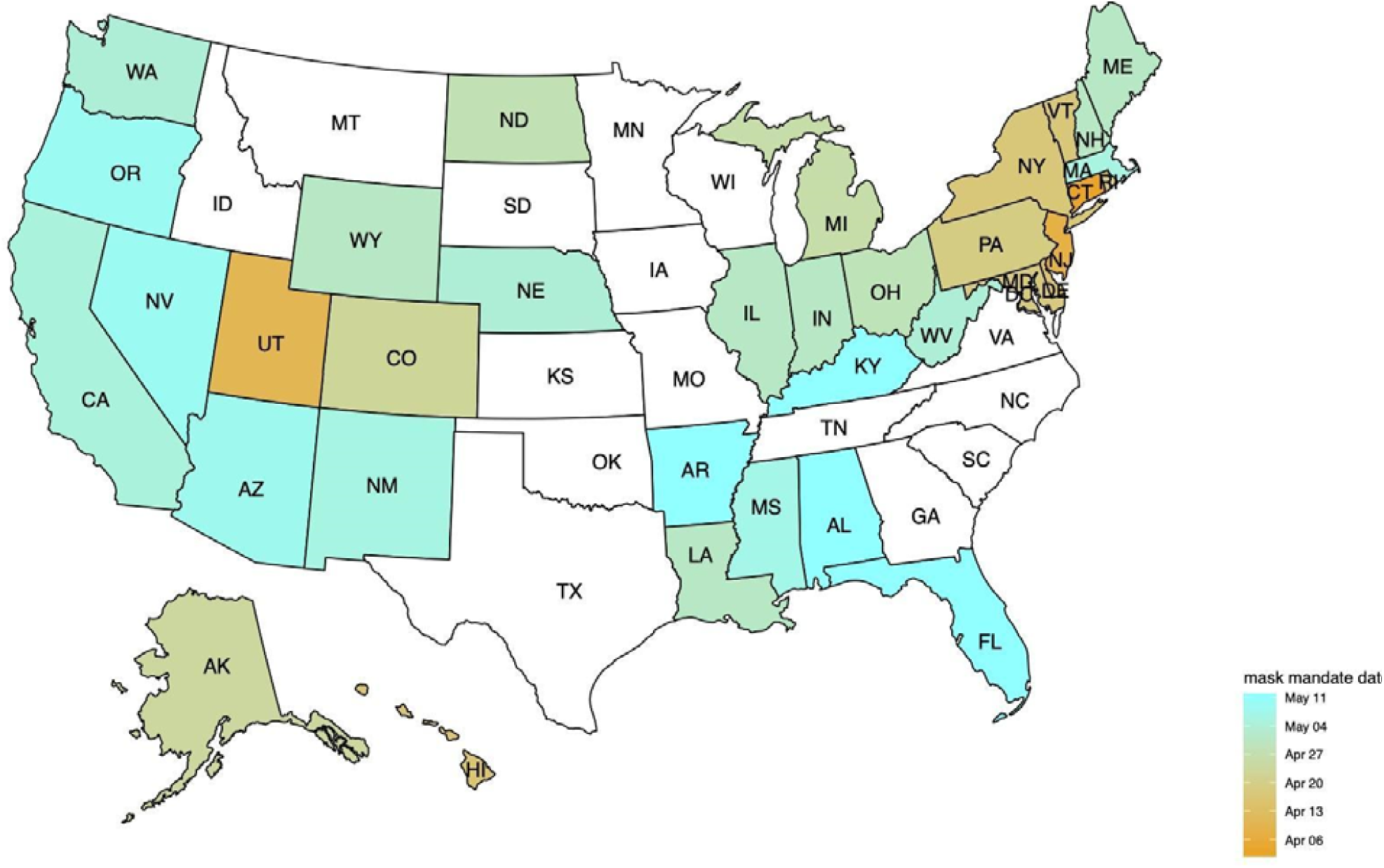
States in the US issuing COVID19’s mandate of mask use orders in April and early May, 2020. 36 states issued the face mask mandates. States in white had not issued a face mask mandat by May 13, 2020.

We use SafeGraph^1^ home dwell time and public visitation data to evaluate the effect that face mask orders had on representative behaviors that could expose individuals to COVID-19 transmission. SafeGraph reports the median home dwell time by census block group, and we produce a device weighted average for each county. We also produce device weighted averages of trip visits to points of interest by 4 digit NAICS code.

### Time at Home

We focus on behavior aligned to two weeks before and after each of the 36 states implemented face mask orders, with a robustness check to a single week window (zero is set at the date of the order for each state). For robustness, we include states that did not issue orders in the control group by using their previous week or two-week data before May 11. For a county *i* in day *t*, we regress time spent at home measured as the device weighted county means of median Census Block Group home dwell time in minutes, *Y_it_*, on the first mask order, *M_it_*. We condition the regression on county-level weather *X_it_*, on reported cases in one’s own county and nationwide, *C_it_* (Yan et al. 2020), a county-specific fixed effect *a_i_*, and a weekday specific fixed effect *w_t_*. Weather variables are constructed by aggregating 4km gridded estimates of maximum and minimum daily temperature, maximum and minimum relative humidity, precipitation amount, surface solar radiation, and wind speed. The two week window in important to capture enough variation to overcome day of the week effects. We cluster standard errors at the state level to account for state-level serial correlation and heteroscedasticity caused by the phase-in orders. The model is specified as

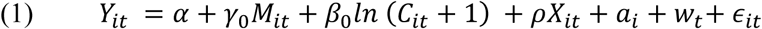

If people attempt to manage infection risk by substituting face mask use for time at home, then we hypothesize that people spend less time at home once they receive a directive to wear masks, *γ*_0_ <0. We include an additional group of policy variables to examine the possible effects of official business reopening in some states. The vector, *B_it_*, includes policy dummies of businesses, restaurants, movie theaters, and gyms being allowed to reopen.

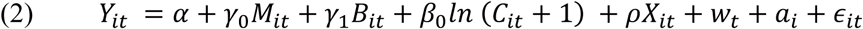

Next, we consider the possibility that individuals have become exhausted with stay-at-home orders (Springborn et al. 2015). We include the log time since the stay at home order, *S_it_*, went into effect. The log specification accounts for the multiplicative nature of this potential effect.

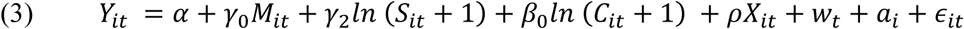

Finally, we investigate possible spillover effects across the state border. We hypothesize that the mandate of mask use issued in a border state, defined as a state bordering the studied county *i*’s state, decrease the dwelling time at home for people in county *i*. The corresponding behavior may reflect a broader level of disinhibition because of spillover from border states, modeled as *bM_it_*. This model can only be estimated over the two-week window because of data limitations.

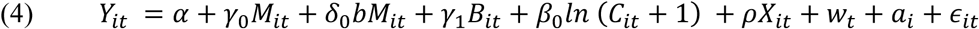

We report results for regressions on the *Y_it_* and on *log*(*Y_it_*) for the above equations. In the supplemental material, we examine pairs of states with and without orders and analyze these pairs with a difference-in-difference design.

### Points of Interest Visitation

If people decrease their time at home, they must go somewhere. It is important to know if they allocate time to relatively high risk or low-risk locations. Benzell, Collis, and Nicolaides (2020) argue that gyms and grocery stores are relatively high risk and hardware stores, sporting goods stores, and general merchandise stores are moderate-risk locations. Conversely, parks may be relatively low-risk locations.

To explore the impact of the mandate of face mask use to site visits, we use points-of-interest (POI) data from March 1 to May 2, 2020, from SafeGraph to examine the change in visits after the face mask mandate.

We aggregate each day *t*’s visits to a site in industry *I* located in county *i, V_Iit_*. Each industry type is analyzed independently. *I* is defined by the first four digits of a locations North American Industry Classification System (NAICS) code. The change in *V_Iit_* is also subject to *c_it_*, the count of devices active in the sample, which varies by county. We regress the county-aggregated visits per device, *ν_nt_ = V_Iit_/c_it_*, on the mask order, *M_it_*. Similar to Equation (1), we condition the regression on county-level weather, *X_it_*, on reported cases in own county and nationwide, *C_it_*, a county fixed effect *a_i_*, and a weekday fixed effect *w_t_*.

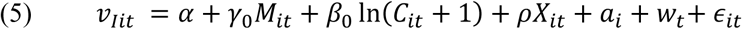

We focus on the sites in the wholesale trade (NAICS sector #41/42), retail trade (NAICS sector #44/45), entertainment and recreation (NAICS sector #71), and accommodation and food services (NAICS sector #72). Consistent with the analysis of time at home, we examine the impact of *M_it_* before and after 14 days of the mask mandate.

We also consider the origin of visitors, which is only available at a weekly aggregate level. We regress the weekly aggregated visits per device, *ν_jw_ = V_jw_/c_iw_* in the point of interest’s county by the visitors home county on a post-mandate dummy *M_jw_*, conditional on the point of interests county-level weekly-average weather, *X_iw_*, state-level fixed effects, *a_s_*, and the site *j*’s industrial group *I_j_·I_j_* is defined as the first 4 digits of its NAICS code.

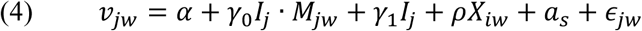

We examine the post-mandate impacts to four groups of visitor-site pairs, i.e., visitors from the same county of the site, outside the county of the site, from the same state of the site, and outside the state of the site.

## Results

We find evidence that masks enable disinhibition behavior and that Americans spend less time at home and more time in moderate to high-risk locations following orders to wear masks.

### Time at home

Americans appear to have reduced time at home following state mandates to wear face masks (Figure 2). Estimates are stable across all specifications and range from a reduction in time from 20-33 minutes or 3-5 percent (Table 1). Business reopening policies do not affect the estimates. This is likely because few states in closed or opened businesses within the window of analysis. However, six do have the same date of reopening as mandatory mask use. Dropping these states does not substantially affect the results. The reported cases at the county and the national levels also do not significantly change the time at home. Yan et al. (2020) find a strong effect of case reports on time at home behavior. However, it is likely that the case effect has largely saturated during the short window of the study. Furthermore, while most counties report new cases, the media focus had shifted during this period.

**Figure 2.**
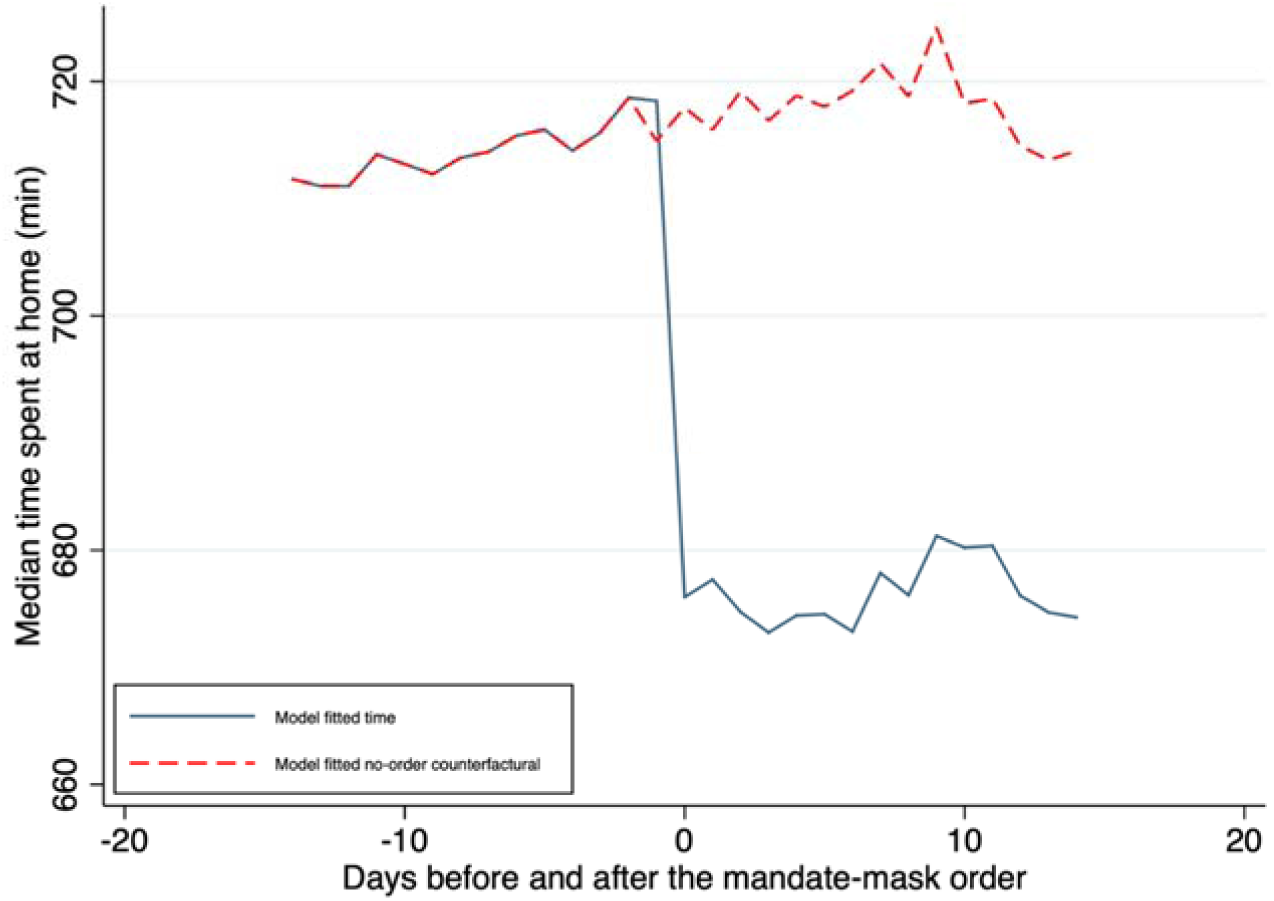
Average time at home before and after COVID-19 face mask mandate.

**Table 1.** Time at home in minutes following COVID19 face mask mandates.

| 14-day window,<br>mandate states | log(y) | y | log(y) | y | log(y) | y | log(y) | y |
| --- | --- | --- | --- | --- | --- | --- | --- | --- |
| Mask mandate | -0.0481***<br>(0.00886) | -32.34***<br>(6.235) | -0.0426***<br>(0.00918) | -29.15***<br>(6.887) | -0.0321**<br>(0.00968) | -19.51**<br>(6.586) | -0.0405***<br>(0.0103) | -25.86***<br>(7.142) |
| Border state's mask<br>mandate |  |  |  |  |  |  | -0.0253*<br>(0.0124) | -21.17*<br>(8.431) |
| Log(days since stay-at-home order issued) |  |  |  |  | -0.0692*<br>(0.0260) | -55.67**<br>(19.20) |  |  |
| Reopen businesses |  |  | -0.00570<br>(0.00983) | -5.967<br>(7.568) |  |  | -0.000490<br>(0.0106) | -1.613<br>(7.787) |
| N | 33981 |  |  |  |  |  |  |  |
| 14-day window, all<br>states | log(y) | y | log(y) | y | log(y) | y | log(y) | y |
| Mask mandate | -0.0499***<br>(0.00929) | -33.71***<br>(6.733) | -0.0489***<br>(0.00992) | -33.19***<br>(7.482) | -0.0357***<br>(0.00928) | -21.46**<br>(6.501) | -0.0466***<br>(0.00946) | -29.21***<br>(6.792) |
| Border state's mask<br>mandate |  |  |  |  |  |  | -0.0179<br>(0.00952) | -18.39**<br>(6.075) |
| Log(days since stay-at-home order issued) |  |  |  |  | -0.0602*<br>(0.0227) | -51.83**<br>(16.80) |  |  |
| Reopen businesses |  |  | 0.0134<br>(0.0130) | 3.284<br>(7.496) |  |  | 0.0176<br>(0.0135) | 7.818<br>(7.654) |
| N | 53533 |  |  |  |  |  |  |  |
| 7-day window, mandate<br>states | log(y) | y | log(y) | y | log(y) | y |  |  |
| Mask mandate | -0.0230*<br>(0.00979) | -15.29*<br>(6.406) | -0.0232*<br>(0.00990) | -16.13*<br>(6.780) | -0.000532<br>(0.00892) | 0.689<br>(5.855) |  |  |
| Log(days since stay-at-home order issued) |  |  |  |  | -0.159***<br>(0.0433) | -113.3***<br>(25.98) |  |  |
| Reopen businesses |  |  | 0.00807<br>(0.0114) | 5.197<br>(7.712) |  |  |  |  |
| N | 19432 |  |  |  |  |  |  |  |
| 7-day window, all states | log(y) | y | log(y) | y | log(y) | y |  |  |
| Mask mandate | -0.0220*<br>(0.0101) | -15.46*<br>(6.965) | -0.0225*<br>(0.0106) | -15.77*<br>(7.286) | 0.00523<br>(0.00936) | 4.559<br>(6.228) |  |  |
| log(days since stay-at-home order issued) |  |  |  |  | -0.176***<br>(0.0480) | -125.1***<br>(28.97) |  |  |
| Reopen businesses |  |  | 0.00450<br>(0.0104) | 2.866<br>(7.087) |  |  |  |  |
| N | 28456 |  |  |  |  |  |  |  |

Clustered standard errors (state level) in parentheses \* p<0.05 \*\* p<0.01 \*\*\* p<0.001
County-level Fixed Effects Model. Week-day fixed effects, county-level weather controls are included.

There is evidence that social distance fatigue is occurring. Time since stay home orders were issued has a greater effect than face mask orders on reducing time at home. Nevertheless, the effect of face masks remains stable. We also find some evidence of policy spillovers and that people are reducing time at home in bordering states that impose face mask orders. However, Americans in a no-policy state do not respond significantly to the bordering states’ mask mandate (Supplement Table 1).

The general results are robust to shortening the window considered to seven days (Table 1). However, the results are less precisely estimated. Furthermore, the effect size is reduced by approximately half. This reduction in effect follows directly from the nature of the model, and the distance between the mean evaluation points. Therefore, the effects are largely consistent with the primary specification. The one exception is that when time since stay-at-home orders were put in place is included, then we do not find an effect of face mask orders. However, the effect of stay-at-home orders is also substantially different than the 14-day specification. We suspect that this is related to day of the week effects.

In the supplement, we provide a difference-in-difference analysis using boundary states. Selecting appropriately paired states is challenging. The paired state analysis does not contradict our primary findings, but is generally ambiguous. Signs mostly conform to the substitution hypothesis but are not significant for the pairs with the strongest evidence of parallel pre-trends. Comparisons involving Wyoming tend to have the opposite sign. We include this analysis for transparency (Ferraro and Shukla 2020).

### Points of Interest Visitation

Americans increased trips to a variety of places in the weeks following the mask mandate (Figure 3 and Table 2). The greatest effects were for restaurants and other eating places, which may include visits for take-out), followed by gas stations, and building supplies. The one shopping location type that did not appear to receive more trips is grocery stores. This may be reasonable since this is one of the few locations open for regular activity throughout the epidemic. Importantly, we look at visits, not time on site. Along with shopping locations, outdoor recreation sites also experienced increase trips. Overall, the increase in out-of-the-house exposure appears to expose people to a mix of sites with different risk characteristics. Nevertheless, most of these sites likely increase transmission risk relative to staying home.

**Figure 3.**
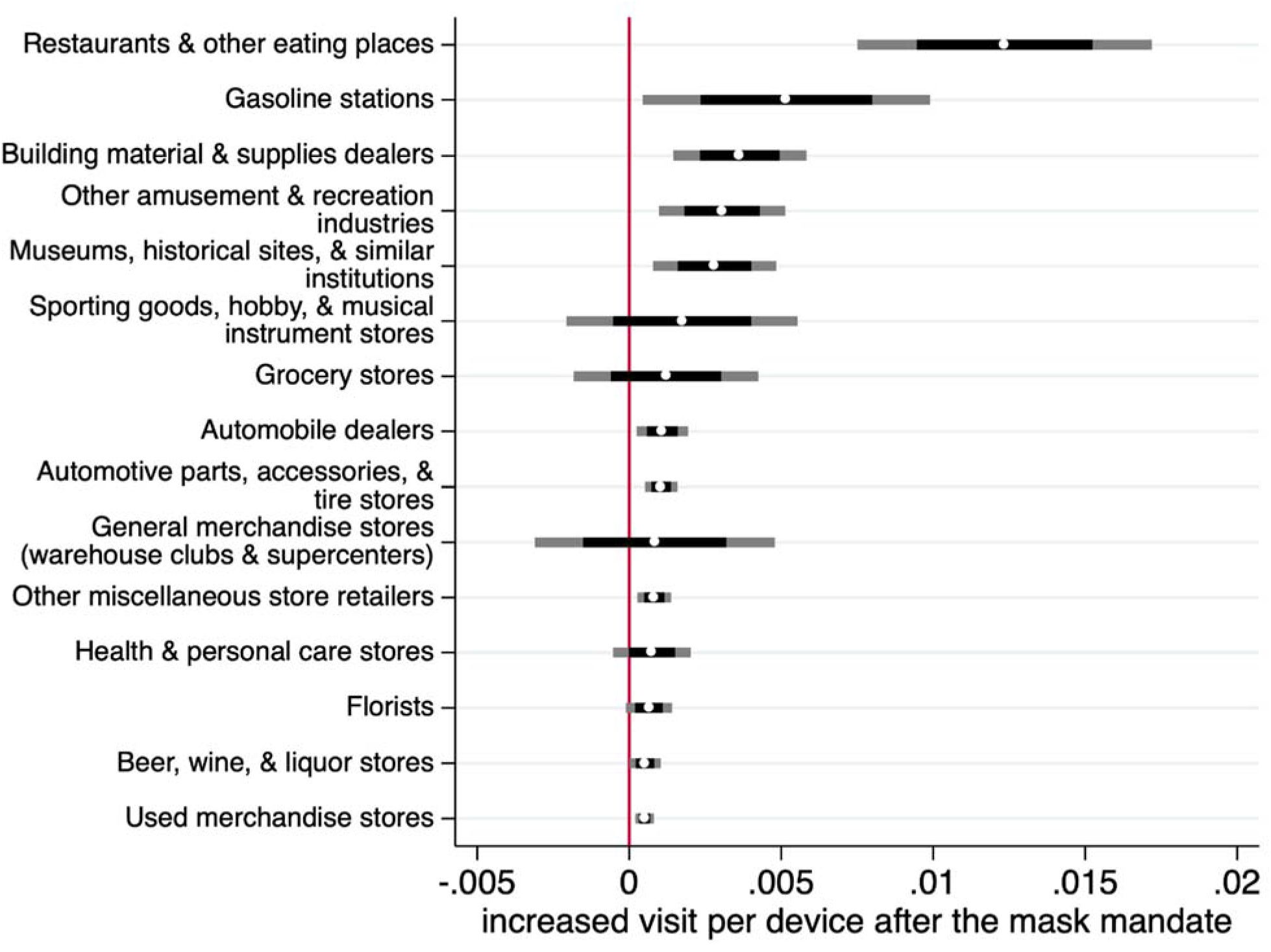
Estimates of increase in visits to a sample of specific types of locations following the face mask order with 95% and 99.85% confidence intervals, full table with additional industries is in the supplemental material (S Table 3). A Bonferroni correction suggests that the 99.85% confidence interval is what should be used conservatively for a 5% probability of a type I error, to address concerns about multiple testing. Parks are included in Museaums, historic sites, & similar instutions. Golf courses are included in Other amusement & recreation industries.

**Table 2.** Increased weekly visit counts per device for a site after the week of COVID19’s mask mandate (broken out by the source of visitors)

| Visitors and POI's location | same county | outside county | outside state |
| --- | --- | --- | --- |
| Wholesale & Retail Sectors |  |  |  |
| Merchandise Stores (Warehouse Clubs & Supercenters)<br>(4523) | 0.00860***<br>(0.00140) | 0.00188***<br>(0.000425) | 0.000773**<br>(0.000255) |
| Lawn and Garden Equipment & Supplies<br>(4442) | 0.00541*<br>(0.00224) | 0.00276*<br>(0.00117) | 0.000811*<br>(0.000324) |
| Department Stores<br>(4522) | 0.00501***<br>(0.000855) | 0.00126***<br>(0.000255) | 0.000319<br>(0.000290) |
| Petroleum and Petroleum Products Wholesalers<br>(4247) | 0.00473***<br>(0.000657) | 0.000743**<br>(0.000255) | 0.000754**<br>(0.000223) |
| Grocery Stores<br>(4451) | 0.00423***<br>(0.000767) | 0.000900**<br>(0.000254) | 0.000321<br>(0.000218) |
| Building Material and Supplies Dealers<br>(4441) | 0.00354***<br>(0.000661) | 0.00152***<br>(0.000311) | 0.000930**<br>(0.000300) |
| Gasoline Stations<br>(4471) | 0.00262**<br>(0.000731) | 0.000590<br>(0.000316) | 0.000235<br>(0.000295) |
| Health and Personal Care Stores<br>(4461) | 0.00219**<br>(0.000693) | 0.000732**<br>(0.000262) | 0.000353<br>(0.000213) |
| Beer, Wine, and Liquor Stores<br>(4453) | 0.00204**<br>(0.000664) | 0.000743**<br>(0.000263) | 0.000373<br>(0.000218) |
| Other Miscellaneous Store Retailers<br>(4539) | 0.00183*<br>(0.000738) | 0.000783**<br>(0.000265) | 0.000311<br>(0.000215) |
| Office Supplies, Stationery, and Gift Stores<br>(4532) | 0.00173*<br>(0.000715) | 0.000653*<br>(0.000286) | 0.000335<br>(0.000206) |
| Specialty Food Stores<br>(4452) | 0.00168*<br>(0.000687) | 0.000696*<br>(0.000291) | 0.000328<br>(0.000280) |
| Sporting Goods/Hobby/Musical Instrument Stores<br>(4511) | 0.00161*<br>(0.000687) | 0.000757**<br>(0.000255) | 0.000504<br>(0.000254) |
| Used Merchandise Stores<br>(4533) | 0.00158*<br>(0.000674) | 0.000670*<br>(0.000275) | 0.000459<br>(0.000228) |
| Clothing Stores<br>(4481) | 0.00129<br>(0.000736) | 0.000639*<br>(0.000288) | 0.000224<br>(0.000231) |
| Book Stores and News Dealers<br>(4512) | 0.00129<br>(0.000791) | 0.000587*<br>(0.000289) | 0.000252<br>(0.000229) |
| Shoe Stores<br>(4482) | 0.00127<br>(0.000754) | 0.000639*<br>(0.000272) | 0.000151<br>(0.000247) |
| Automobile Dealers<br>(4411) | 0.00116<br>(0.000741) | 0.000546<br>(0.000284) | 0.000173<br>(0.000229) |
| Recreation & Food Service Sectors |  |  |  |
| Restaurants and Other Eating Places<br>(7225) | 0.00203**<br>(0.000668) | 0.000757**<br>(0.000261) | 0.000355<br>(0.000209) |
| Performing Arts Companies<br>(7111) | 0.00200*<br>(0.000796) | 0.000632*<br>(0.000242) | -0.0000618<br>(0.000181) |
| Gambling Industries | 0.00178** | 0.000714* | 0.000656 |
| (7132) | (0.000652) | (0.000341) | (0.000366) |
| Traveler Accommodation | 0.00166* | 0.000532 | 0.000138 |
| (7211) | (0.000669) | (0.000332) | (0.000305) |
| Other Amusement and Recreation Industries | 0.00147* | 0.000610* | 0.000222 |
| (7139) | (0.000710) | (0.000282) | (0.000219) |
| Drinking Places (Alcoholic Beverages) | 0.00144* | 0.000648* | 0.000232 |
| (7224) | (0.000696) | (0.000276) | (0.000231) |
| Amusement Parks and Arcades | 0.00137* | 0.000516 | 0.000251 |
| (7131) | (0.000671) | (0.000335) | (0.000247) |
| RV (Recreational Vehicle) Parks and Recreational<br>Camps | 0.00683 | 0.000260 | -0.000101 |
| (7212) | (0.00434) | (0.000364) | (0.000445) |
| Museums, Historical Sites, and Similar Institutions | 0.00146 | 0.000620 | 0.000364 |
| (7121) | (0.000735) | (0.000328) | (0.000272) |
| Spectator Sports | 0.000598 | 0.000382 | 0.000129 |
| (7112) | (0.000866) | (0.000406) | (0.000293) |
| N | 4114914 | 3576905 | 987872 |

Clustered standard errors (state level) in parentheses \* $p < 0.05$ \*\* $p < 0.01$ \*\*\* $p < 0.001$
Coefficients of the post-mandate dummy with industrial group interactions are reported. County-level weather controls are included.

By aggregating trips across the week it is possible to parse the visitor’s census block group origin (Table 3). It is noticeable that certain industries have a significant increase in visits from visitors outside of the location’s state following the face mask mandate. For example, visits from out of state visitors to warehouse clubs and supercenters increased by 0.0008 trip per device, or approximately 4.6 trips. We also note that using the weekly visits we do find an increase in trips to grocery stores.

### Addressing potential concerns

It is possible that distancing fatigue or cabin fever coincided with the implementation of mask orders. However, if this were the case, then we would expect that behavior driven by distancing fatigue would be gradual and would not systematically correspond to mask orders implemented differentially over space and time. This is difficult to test. We attempt to control for distancing fatigue with the log time since stay at home orders went into effect. Using the fourteen-day window, the results are robust. However, it is likely that there is some distancing fatigue happening, which makes it difficult to measure the disinhibition from the face mask wearing effect separately from distancing fatigue. Nevertheless, the disinhibition effect appears robust.

Alternatively, one could be concerned the governors are simply running to the front of the crowd. The assumption would be that governors know people will start going out. Therefore, they instruct face mask use to signal it is ok to start going out. If this were the case, it is unlikely that there would be a systematic break, and a substantial number of governors are relaxing stay at home orders directly. Furthermore, such a concern simultaneously gives the governors a large amount of credit for political astuteness and treats their actions with a high degree of cynicism. Both assumptions seem unmerited in the fog of the COVID-19 crisis.

Finally, we analyze the effect of the mask orders defined by the implementation date and not the announcement. It is possible that Governors’ policy announcements or earlier CDC announcements signaled to the public that it is safe to resume public interaction. Such anticipatory behavior would attenuate our estimates because the pre-policy period would be contaminated by the behavioral response we associate with the face mask orders. Still, we find a robust decrease in time spent and home and a robust increase in trips to public places following the implementation of face mask requirements.

## Discussion

Americans are increasing visitation to public locations and reducing their time spent at home. This happens even as COVID-19 cases continue to rise in most of the United States. Our results suggest that mask orders provide a sense of protection, leading people to substitute face mask wearing for other non-pharmaceutical interventions like avoiding time in public. The net effect of these behaviors on public health outcomes depends on the relative effectiveness of masks and other behaviors in reducing transmission.

Evidence suggests that staying home effectively reduced transmission. One concern with that conclusion is that it is easier to observe staying home behavior than hand-washing and face mask-wearing. Yet, the evidence of the effectiveness of face mask use by the general public on disease transmission is less conclusive. Furthermore, for face masks to be effective, they must be used correctly. This includes having a tight seal, which requires things like a clean shave, having one’s nose in the mask, and leaving the mask on while talking to someone. One need only look at images of mask-wearing in public to conclude that a non-trivial share of the mask wearers are not wearing them correctly or that the masks themselves are of questionable quality (i.e., bandannas). It is certainly possible that misused masks do not increase transmission, and may reduce it, holding other behaviors constant. The challenge is that the introduction of the wide use of face masks appears to alter those other behaviors.

Our results suggest that wearing a mask needs to be as effective as all individuals in society reducing their time out of the house by approximately 4 percent or 20-30 minutes. Bayham et al. (2015) found that voluntary behavioral change of a similar magnitude reduced swine flu cases on the order of 10 percent. This is likely a lower bound for the value of an additional 20-30 minutes at home for reducing COVID-19 cases. Are face masks that effective? Our results provide a benchmark for future testing of the effectiveness of face masks in the general public.

Messaging around behavioral interventions needs to be done carefully. Masks have been introduced. Our results should not be used as a justification for discouraging face mask use. Rather, extreme care must be taken when suggesting new behaviors that may be helpful in order to avoid replacing behaviors that are known to be helpful. The message to wear a face mask in public is at least suggestive that it is safer to resume public interactions with a mask. However, time in public is still riskier than time at home and can still enable transmission from asymptomatic individuals.

Concerns about behavioral substitution are sometimes treated as curiosities. Requiring seat belts likely outweighs the damage from riskier driving and energy efficiency reduces carbon emissions even if it leads to more device use (Gillingham et al. 2013). Even encouraging condom use has likely prevented more cases of HIV, then the risk compensation generated. However, in the case of the highly contagious pathogens SAR-COV-2, which causes COVID-19, a small amount of risk compensation behavior could lead to an exponential increase in cases. The difference between a few trips with a mask and staying home, spread across the entire population could be the difference between the reproductive rate of the pathogen (R(t)) exceeding one, and renewed exponential growth, and a reproductive rate less than one and containing the epidemic. If people must go out, then it is probably advisable to wear a mask.

However, the fact that people now own masks is likely making them more likely to even consider a trip to a warehouse club, a home improvement store, or a liquor store. Ultimately, these marginal trips could make the epidemic more difficult to bring under control.

## Data Availability

Data can be acquired easily from SafeGraph and other public sources.

## Acknowledgements

Funding: YY and EPF are supported by the Knobloch Family Foundation.

## JEL Classification Codes

I18, I12, H4,

## Supplementary Materials

In this section, we provide three different robustness checks. S Table 1 reports Americans’ time at home response in a state without a face mask mandate in the previous one to three weeks. The mandates from the bordering states seem not to influence the response in the short run. This also provides a baseline for the effect comparison to states with the mandates.

We further explore the heterogeneous behavioral changes between states to show a more nuanced time response under the mask mandate. We find 39 bordered pairs of states. These include 27 with one state having a mask mandate before May 11^th^ and the other state never has a mandate before May 11^th^. For the other 12 pairs mask mandates are separated by at least 14 days. For each pair, we apply a difference-in-difference analysis to examine the effect of the mandate. The results are in S Table 2. The majority of the pairs exhibit the expected sign suggesting that individuals substitute face masks for staying home. However, many of the estimates are imprecise. There are pairs where the sign suggests wearing a face mask is a complement to staying home. This seems unlikely and most of these pairings involve Wyoming, suggesting there may be an important unobservable associated with that state.

We order the coefficients in S Table 2 of the key results by the p-value of the pre-policy’s parallel trend assumption. Difference-in-difference assumes parallel pre-trends. We compare the coefficient by regressing time at home for 14 days before the face mask policy in state 1 and the corresponding coefficient in state 2 using an F-test. The lower the p-value the more questionable the parallel pre-trends assumption is, which is equivalent to making a type II error when relying on the parallel trends assumptions. A low p-value would lead us to reject the null that the pre-trends are indeed parallel.

Supplement Table 3 provides a table version of Figure 3 with additional sectors included.

**S Table 1.** Time at home (min) under COVID19’s mandate of mask use for no-mandate states.

|  | previous 1 week |  | previous 2 weeks |  | previous 3 weeks |  |
| --- | --- | --- | --- | --- | --- | --- |
|  | log(y) | y | log(y) | y | log(y) | y |
| Border state's mask mandate | -0.00578<br>(0.0144) | -3.445<br>(8.857) | 0.0201<br>(0.0146) | 4.436<br>(6.191) | -0.00504<br>(0.00871) | -9.447*<br>(4.164) |
| log(county case) | 0.00732<br>(0.0169) | -0.690<br>(6.931) | 0.00598<br>(0.0113) | 8.814<br>(5.150) | -0.0127<br>(0.00701) | -6.437<br>(3.762) |
| log(national case) | -0.00402*<br>(0.00165) | -1.722*<br>(0.753) | 0.000474<br>(0.000764) | 0.129<br>(0.377) | 0.000692<br>(0.000620) | 0.364<br>(0.356) |
| Re-open businesses | 0.0234*<br>(0.00882) | 15.57*<br>(5.450) | 0.0314<br>(0.0160) | 15.72*<br>(7.265) | 0.00931<br>(0.0128) | 2.459<br>(5.880) |
| N | 9024 |  | 19552 |  | 30080 |  |

Clustered standard errors (state level) in parentheses \* p<0.05 \*\* p<0.01 \*\*\* p<0.001
County-level Fixed Effects Model. Week-day fixed effects, county-level weather controls are included.

**S Table 2.** Comparison of time at home (min) under COVID19’s mandate of mask use between bordered states (the left is the ordered-state or first ordered-state, N is for total observations)

| pair of states | logy | y | p-val for parallel trend test | (cont'd) | logy | y | p-val for parallel trend test |
| --- | --- | --- | --- | --- | --- | --- | --- |
| DC-VA | -0.0364 | -21.36 | 0.895 | ND-MT | -0.0160 | -16.75** | 0.492 |
| (N=3770) | (0.0264) | (16.84) |  | (N=2834) | (0.0172) | (6.367) |  |
| WA-ID | 0.0104 | -16.23** | 0.887 | UT-ID | -0.00630 | 0.0320 | 0.442 |
| (N=1660) | (0.0256) | (5.711) |  | (N=2117) | (0.0201) | (5.691) |  |
| NM-TX | -0.0198 | 3.487 | 0.874 | NM-OK | -0.0269 | 2.008 | 0.261 |
| (N=5166) | (0.0189) | (5.987) |  | (N=1980) | (0.0212) | (4.720) |  |
| WY-SD | 0.0922** | 42.24*** | 0.840 | CO-OK | -0.0683*** | -35.23*** | 0.167 |
| (N=2047) | (0.0293) | (11.14) |  | (N=4089) | (0.00744) | (3.871) |  |
| NV-ID | -0.155 | -38.68 | 0.817 | PA-WV | -0.0588*** | -45.73*** | 0.154 |
| (N=915) | (0.102) | (19.97) |  | (N=3538) | (0.00416) | (2.692) |  |
| CT-RI | 0.0127 | 9.259 | 0.803 | UT-NV | -0.0128 | -4.885 | 0.152 |
| (N=377) | (0.0139) | (9.534) |  | (N=1334) | (0.0187) | (6.361) |  |
| RI-MA | -0.00999 | -6.950 | 0.803 | NE-KS | -0.0548*** | -11.56* | 0.131 |
| (N=551) | (0.0113) | (8.804) |  | (N=3960) | (0.0137) | (4.563) |  |
| WY-MT | 0.0546* | 33.43*** | 0.800 | UT-NM | -0.0177 | -9.972 | 0.108 |
| (N=1817) | (0.0238) | (9.056) |  | (N=1798) | (0.0189) | (5.995) |  |
| WY-ID | 0.00758 | 25.88*** | 0.725 | MI-WI | -0.0705*** | -51.36*** | 0.107 |
| (N=1541) | (0.0315) | (7.225) |  | (N=4340) | (0.00485) | (3.014) |  |
| ND-SD | -0.0139 | -7.182 | 0.724 | UT-AZ | -0.0247* | -10.30 | 0.096 |
| (N=3094) | (0.0142) | (5.303) |  | (N=1276) | (0.0109) | (5.811) |  |
| WV-VA | -0.00681 | -5.318 | 0.701 | ND-MN | -0.0296** | -20.14*** | 0.091 |
| (N=3680) | (0.00583) | (3.376) |  | (N=3640) | (0.0108) | (4.541) |  |
| VT-MA | -0.0647*** | -40.58*** | 0.621 | LA-TX | 0.000713 | -3.128 | 0.063 |
| (N=812) | (0.00821) | (5.625) |  | (N=7314) | (0.00773) | (3.504) |  |
| NY-MA | -0.0703*** | -53.14*** | 0.607 | IL-MO | -0.00774* | -8.107*** | 0.045 |
| (N=2204) | (0.00422) | (2.947) |  | (N=4991) | (0.00304) | (2.251) |  |
| MD-VA | -0.0159** | -15.13*** | 0.607 | IL-IA | -0.00221 | -4.265* | 0.036 |
| (N=4437) | (0.00587) | (3.745) |  | (N=4623) | (0.00286) | (2.129) |  |
| NE-SD | -0.0671*** | -18.11*** | 0.595 | NE-MO | -0.0531*** | -11.64** | 0.029 |
| (N=3180) | (0.0161) | (5.005) |  | (N=4160) | (0.0131) | (4.157) |  |
| MD-WV | -0.0164* | -15.44** | 0.587 | IL-WI | -0.000831 | -3.796 | 0.011 |
| (N=2291) | (0.00761) | (4.826) |  | (N=4002) | (0.00374) | (2.480) |  |
| OR-ID | -0.0590 | -22.80 | 0.580 | NE-IA | -0.0524*** | -11.67** | 0.009 |
| (N=1200) | (0.0620) | (13.28) |  | (N=3840) | (0.0122) | (3.753) |  |
| CT-MA | -0.0549*** | -21.79** | 0.578 | UT-WY | -0.0395*** | -17.44** | 0.005 |
| (N=638) | (0.0111) | (7.097) |  | (N=1508) | (0.00988) | (5.630) |  |
| CO-KS | -0.0731*** | -42.22*** | 0.557 | MS-TN | 0.0156** | 9.368** | 0.000 |
| (N=4901) | (0.00764) | (4.134) |  | (N=3009) | (0.00556) | (3.518) |  |
| NJ-DE | 0.00446 | 12.81** | 0.493 |  |  |  |  |
| (N=696) | (0.00532) | (4.596) |  |  |  |  |  |

Clustered standard errors (state level) in parentheses \* p<0.05 \*\* p<0.01 \*\*\* p<0.001

**S Table 3.** County-level visit counts per device for a retail, wholesale, or recreation site under the COVID19’s mask mandate in the site’s state (14-day before and after the mandate comparison)

| Increased visit per device to a site in the Industry of: | Coeff. of mask mandate | SE |
| --- | --- | --- |
| Restaurants and Other Eating Places | 0.0172*** | (0.00179) |
| Gasoline Stations | 0.00654*** | (0.00134) |
| Building Material and Supplies Dealers | 0.00496*** | (0.000665) |
| Other Amusement and Recreation Industries | 0.00332*** | (0.000566) |
| Museums, Historical Sites, and Similar Institutions | 0.00274** | (0.000813) |
| General Merchandise Stores: Warehouse Clubs and Supercenters | 0.00221* | (0.000828) |
| Sporting Goods, Hobby, and Musical Instrument Stores | 0.00189* | (0.000858) |
| Automobile Dealers | 0.00133*** | (0.000268) |
| Grocery Stores | 0.00129 | (0.000850) |
| Automotive Parts, Accessories, and Tire Stores | 0.00122*** | (0.000149) |
| Other Miscellaneous Store Retailers | 0.00101*** | (0.000134) |
| Health and Personal Care Stores | 0.000930** | (0.000285) |
| Florists | 0.000715*** | (0.000136) |
| Beer, Wine, and Liquor Stores | 0.000628*** | (0.000113) |
| Used Merchandise Stores | 0.000616*** | (0.0000882) |
| Office Supplies, Stationery, and Gift Stores | 0.000495*** | (0.000120) |
| Department Stores | 0.000460*** | (0.000122) |
| Specialty Food Stores | 0.000364*** | (0.0000871) |
| Furniture Stores | 0.000362*** | (0.0000595) |
| Clothing Stores | 0.000303*** | (0.0000731) |
| Electronics and Appliance Stores | 0.000278** | (0.0000799) |
| Jewelry, Luggage, and Leather Goods Stores | 0.000226*** | (0.0000637) |
| Home Furnishings Stores | 0.000221*** | (0.0000414) |
| Book Stores and News Dealers | 0.000189*** | (0.0000472) |
| Gambling Industries | 0.000132 | (0.0000704) |
| Machinery, Equipment, and Supplies Merchant Wholesalers | 0.0000645* | (0.0000256) |
| Shoe Stores | 0.0000516* | (0.0000216) |
| Amusement Parks and Arcades | 0.0000493 | (0.0000274) |
| Miscellaneous Durable Goods Merchant Wholesalers | 0.0000489* | (0.0000238) |
| Lumber and Other Construction Materials Merchant Wholesalers | 0.0000310* | (0.0000135) |
| Drugs and Druggists' Sundries Merchant Wholesalers | 0.0000283 | (0.0000268) |
| Drinking Places (Alcoholic Beverages) | 0.0000244 | (0.0000516) |
| Chemical and Allied Products Merchant Wholesalers | 0.0000175** | (0.00000628) |
| N | 33483 |  |

Clustered standard errors (state level) in parentheses \* p<0.05 \*\* p<0.01 \*\*\* p<0.001
County-level Fixed Effects Model. Week-day fixed effects, county-level weather controls are included.

## Footnotes

1 SafeGraph is a data company that aggregates anonymized location data from numerous applications in order to provide insights about physical places. To enhance privacy, SafeGraph excludes census block group information if fewer than five devices visited an establishment in a month from a given census block group (https://docs.safegraph.com/docs/social-distancing-metrics).

## Notes

### Competing Interest Statement

The authors have declared no competing interest.

### Author Declarations

IRB not required

